# Is Hydroxychloroquine Safe During Pregnancy? Observations from Penn Medicine

**DOI:** 10.1101/2020.04.29.20085621

**Authors:** Lena Davidson, Silvia P. Canelón, Mary Regina Boland

**Author notes:** **Corresponding Author:** Mary Regina Boland, PhD, Assistant Professor of Informatics, Perelman School of Medicine, University of Pennsylvania, 423 Guardian Drive, 421 Blockley Hall, Philadelphia, PA 19104.

## Abstract

A novel strain of coronavirus appeared in December 2019. Over the next few months, this novel coronavirus spread throughout the world, being declared a pandemic by the World Health Organization on March 11, 2020. As of this writing (March 28, 2020) over one hundred thousand individuals in the United States of America were confirmed cases. One way of treating the associated disease, COVID-19, is to reuse existing FDA-approved medications. One medication that has shown promise is hydroxychloroquine (HCQ). However, the utility and safety of HCQ among pregnant COVID-19 patients remains a concern.

---

A recent open-label non-randomized clinical trial of HCQ and azithromycin as treatment for COVID-19 excluded pregnant and breastfeeding patients from the study[1]. HCQ is considered a Category C medication, indicating that it remains unknown what effect the drug will have on the fetus.

There is evidence that HCQ may be safe during pregnancy, with previous research finding no increased risk of, prematurity, fetal death, retinopathy, low birth weight, stillbirth, or congenital defects[2–4]. However, there is also evidence that HCQ could result in fetal harm with a recent meta-analysis finding an association between HCQ use and spontaneous abortion^3^ and small for gestational age[5]. Therefore, clinicians may be faced with the difficult decision of deciding whether to treat their pregnant COVID-19 patients with HCQ.

At Penn Medicine, we have pregnancy outcomes following 63,334 deliveries between 2010 and 2017 with the following outcomes annotated: Caesarean section delivery, preterm birth, multiple birth (e.g., twins, triplets) and stillbirth. We assessed whether there was an increased risk of any of these four outcomes following exposure to HCQ during pregnancy where exposure was determined to occur between 280 days (i.e., 40 weeks) and up to 1 day prior to the date of delivery. The Institutional Review Board at the University of Pennsylvania approved this study. We found that 28 deliveries had documented HCQ exposure, results shown in **Figure**. We did not find increased risk for any of the four measured outcomes (p>0.05): Caesarean Section, Preterm birth, Multiple birth, and Stillbirth. We also expanded our search to include all quine drugs (i.e., HCQ, primaquine, mefloquine, chloroquine phosphate) and found the same negative result (i.e., no increased risk).

Our findings coupled with some of those in the literature[2–4] suggest that HCQ may not adversely affect fetal outcomes when taking HCQ during pregnancy. Therefore, clinicians may consider including pregnant patients with COVID-19 in clinical trials of HCQ when warranted.

## Data Availability

This is a retrospective analysis of existing clinical data - therefore, these data are not available for use directly.

## FUNDING

This work was funded by the generous support of the University of Pennsylvania, Perelman School of Medicine.

**IRB APPROVAL #828000**

---

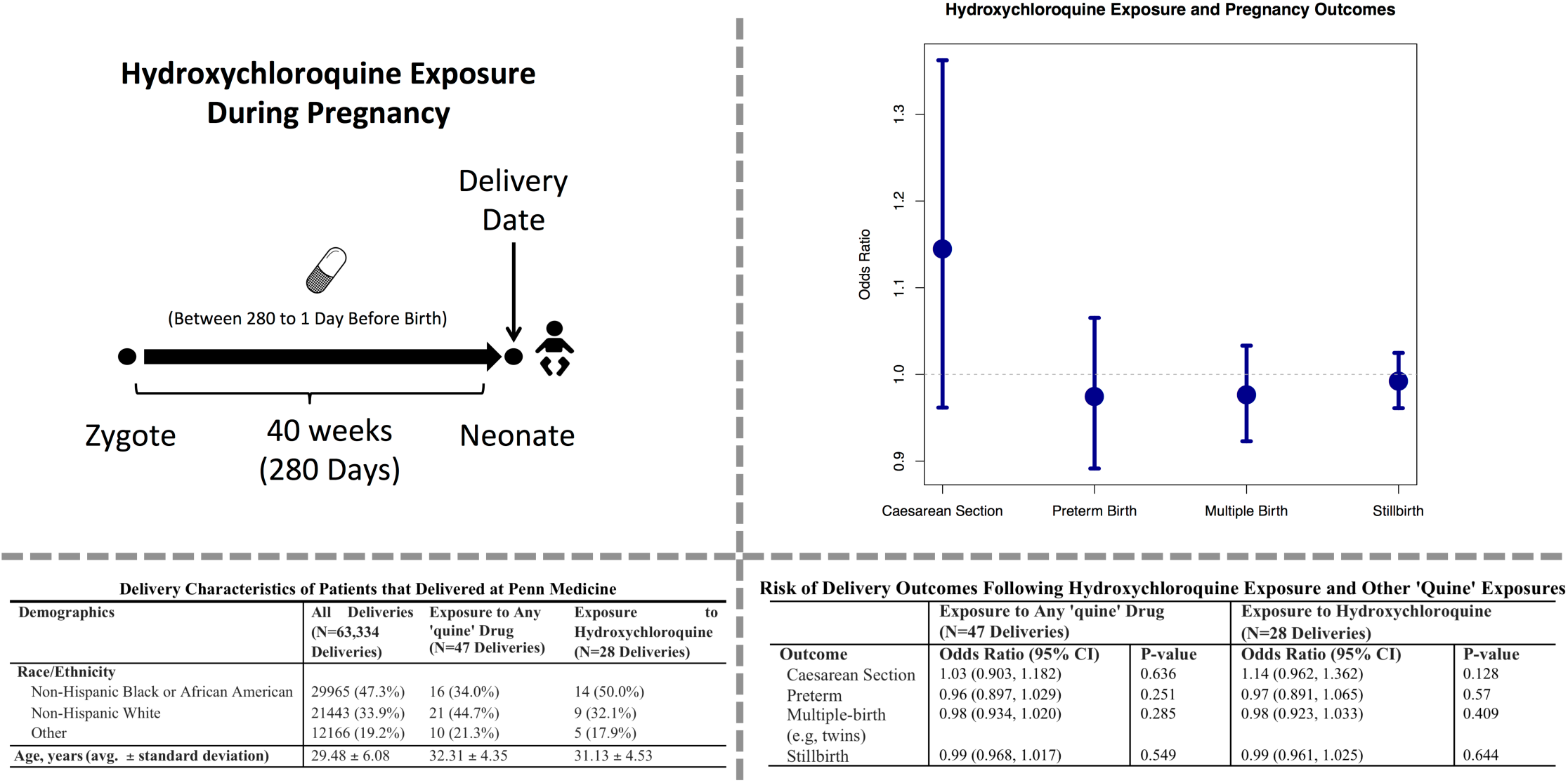
**Figure. Hydroxychloroquine Exposure and Four Delivery Outcomes (Caesarean Section, Preterm Birth, Multiple Birth, Stillbirth) Using PennMedicine Data from 28 Pregnancies**

